# Helping Underdeveloped Lungs with Cells (HULC)-2 Mesenchymal Stromal Cells in extreme preterm infants at risk of developing Bronchopulmonary Dysplasia – A Phase 2 Multi-Centre Double Blind Randomized Controlled Trial Study Protocol

**DOI:** 10.64898/2025.12.11.25337743

**Authors:** Laurent Renesme, Emanuela Ferretti, Chantal Horth, Rebecca Grimwood, Liam Da Sylva, Viviane Olson, Chanele Cyr-Depauw, Daniel Freund, Mario Rüdiger, Marius A. Möbius, Samantha Hodgins, Saad Khan, David Courtman, Dean Fergusson, Bernard Thébaud

## Abstract

**Introduction:** Bronchopulmonary dysplasia (BPD) remains a major complication among extremely low gestational age (ELGA) infants, with long-term respiratory and neurodevelopmental consequences. Despite advances in neonatal care, effective therapies to prevent BPD are lacking. Mesenchymal stromal cells (MSC), particularly those derived from umbilical cord (UC-MSC), offer promise due to their pleiotropic effects. Preclinical and early-phase clinical studies have demonstrated safety and potential efficacy of MSC in neonatal lung injury. The HULC-2 trial aims to evaluate whether multiple intravenous doses of human allogenic UC-MSC can reduce mechanical ventilation duration and improve the respiratory outcome in ELGA infants at high risk of developing BPD.

**Methods and Analysis:** HULC-2 is a multicenter, double-blind, randomized controlled Phase II trial conducted in Canadian Neonatal Intensive Care Units. ELGA infants (gestational age <28 weeks) who remain ventilator-dependent between 4–14 days of life will be randomized to receive either three weekly intravenous doses of UC-MSC (10×10^6 cells/kg/dose) or a sham procedure. The primary outcome is ventilation-free days (VFDs) at 120 days post-randomization, accounting for mortality. Secondary outcomes include cell administration safety, respiratory and neurodevelopmental outcomes, and complications of prematurity. A total of 168 participants will be enrolled to detect a clinically meaningful difference in VFDs.

**Ethics and Dissemination:** Ethics approval has been obtained, and the trial is registered on ClinicalTrials.gov. Results will be disseminated via peer-reviewed publications, conferences, and public engagement platforms. Parent partners are actively involved in study design and dissemination to ensure relevance and transparency.

**Strengths and limitations of this study:**

- Ventilation-free day (VFD) is a clinically relevant primary outcome compared to BPD, as prolonged mechanical ventilation in preterm infants is directly associated with increased risks of mortality, neurodevelopmental impairment, and other complications, making VFDs a more sensitive and meaningful measure of both survival and recovery
- Use of a multiple-dose regimen and fresh cell product to optimize the cell’s therapeutic potential
- Cell product was tested in a large animal model of BPD
- Parents’ involvement in the trial design and development to deliver meaningful research that benefits patients and increase study acceptance among parents.
- A sham procedure was chosen over a placebo based on parent feedback, which increased the risk of unblinding, but strategies to mitigate that risk were developed.

## Introduction

Among preterm infants, those born at extremely low gestational age (or ELGA, born before 28 weeks of gestation) face the highest risk of severe complications, particularly in lung and brain development (1–3). Bronchopulmonary dysplasia (BPD) is a chronic lung disease affecting preterm infants, with an incidence reaching up to 45% in the ELGA population (4,5). BPD arises from impaired lung development due to prematurity and the intensive treatments required to support these fragile infants, such as mechanical ventilation, which subjects their developing lungs to harmful factors, including oxidative stress and inflammation. These factors disrupt normal lung growth and contribute to the development of BPD. Despite advances in perinatal care, preventing lung injury remains challenging, with the incidence of BPD either stagnating or rising (6–8). BPD has lifelong respiratory and neuro-developmental consequences beyond childhood, resulting in increased healthcare costs (5,9,10). Reports indicate arrested alveolar growth in older children (11), lower cognitive development and quality of life scores at 10 years of age (12), early-onset emphysema (13), and pulmonary hypertension (14) in young adults who had BPD. Effective interventions at this stage would provide exceptional value as they would prevent lifelong ailments. Currently, there is no cure for BPD. Management of BPD is primarily supportive, including optimization of positive-pressure ventilation, nutrition, post-natal steroids, and diuretics

(15). Given the multifactorial nature of BPD, traditional pharmacological therapies targeting a single pathway are unlikely to significantly improve outcomes (16). Therefore, there is a clear need for more effective and safe treatments that increase survival without morbidity, promote lung growth and development, and prevent further lung injury.

Mesenchymal stromal cells (MSCs) represent a promising therapy for BPD due to their pleiotropic effects. Preclinical studies have shown that MSCs prevent lung injury in animal models of BPD. Proof of concept studies suggested that bone marrow-derived MSCs prevent impaired alveolar development in neonatal rodent BPD models (17,18). We also demonstrated that airway delivery of human umbilical cord blood-derived MSCs and cord tissue-derived perivascular cells preserves alveolar and lung vascular growth, restores lung function and structure after established lung injury, and results in long-term therapeutic benefit without adverse effects or tumor formation (19). Additionally, this work demonstrated the low engraftment potential of MSCs. Contrary to the original hypotheses, no evidence exists that MSCs can engraft in tissues and persist. Increasing evidence suggests that MSCs exert their beneficial effects through paracrine activity and cell-to-cell contact, explaining their pleiotropic effects, such as anti-inflammatory, anti-fibrotic, and anti-oxidative properties (20–22). A preclinical systematic review on MSCs in experimental neonatal lung injury, while highlighting several shortcomings, supported these findings (23) and provides further biological plausibility for the use of MSCs to prevent BPD. Furthermore, preclinical studies in experimental neonatal lung injury (24,25) and in hypoxic-ischemic brain injury or after intraventricular hemorrhage (26,27) suggest that MSCs have a neuroprotective effect.

Clinical studies using MSCs, including in preterm infants, demonstrate a favorable safety profile. Human allogenic MSCs have been extensively tested in adult clinical trials, and multiple systematic reviews have confirmed their clinical safety profile with no reports of immunogenic rejection or other adverse effects other than transient fever (28–30). A systematic review focusing on clinical trials using umbilical cord-derived MSCs (UC-MSCs) revealed no long-term adverse effects, tumor formation, or cell rejection (28). A more recent systematic review confirmed the safety profile of intravenous (IV) administration of MSCs, with an increased risk of fever but not acute infusion toxicity, infection, thrombotic/embolic events, death, or malignancy (29). Lastly, five clinical studies, including our Phase I study (HULC-1), have evaluated the safety of MSC administration in ELGA preterm infants. No dose-limiting toxicity or serious adverse events were reported, demonstrating the safety and feasibility of MSC therapy in preterm infants (31–34).

Our trial hypothesis is that multiple IV doses of human allogenic UC-MSCs given during the first weeks of life will reduce the duration of mechanical ventilation and improve overall survival in ELGA preterm infants at high risk of developing BPD. This study is designed as a Phase II multicenter randomized controlled trial to evaluate the efficacy and safety of UC-MSC therapy in this vulnerable population.

## Methods and analysis

### Study Design

HULC-2 is a phase II multicenter double-blind randomized controlled trial conducted in tertiary-level neonatal intensive care units (NICUs) across Canada. The study flow diagram is presented in Figure 1.

**Figure 1.**
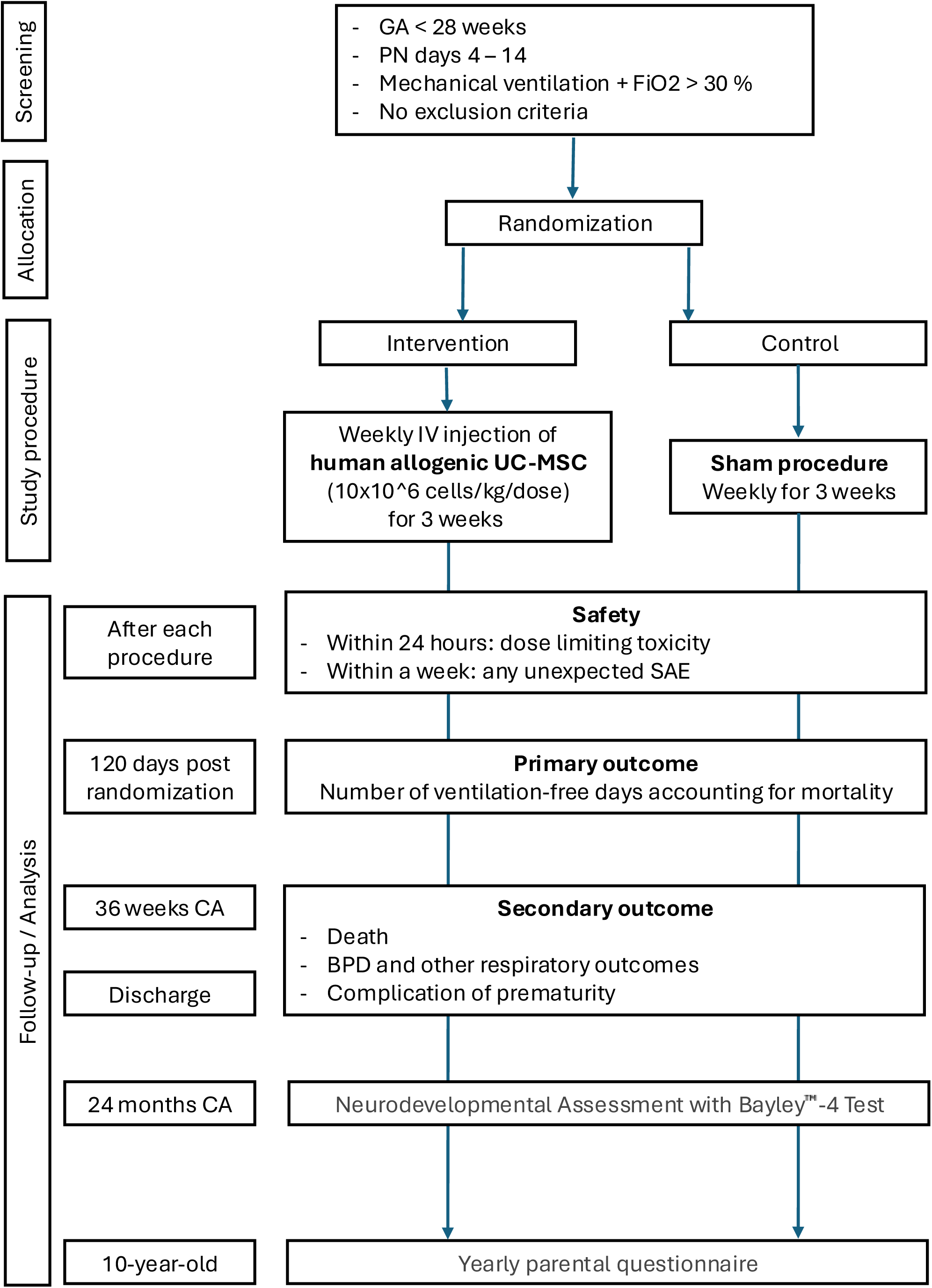
Study flow diagram. GA: Gestational age, PN: postnatal, FiO2: Fraction of inspired oxygen, IV: intravenous, UC-MSC: umbilical cord-derived mesenchymal stromal cell, SAE: serious adverse event, CA: corrected age, BPD: bronchopulmonary dysplasia

### Study Research questions

Our primary research question, developed using the PICO framework (Population, Intervention, Control and Outcomes), is the following: in extremely low gestational age preterm infants on mechanical ventilation, does the administration of multiple IV doses of human allogenic UC-MSCs (1 weekly dose of 10×10^6^ cell/kg for 3 weeks), compared to no UC-MSC injection (Control group with Sham procedure once weekly for 3 weeks), increase the number of mechanical ventilation-free days accounting for mortality? Secondary study questions include confirming the safety profile of IV administration of human allogenic UC-MSC in this population and whether the administration of multiple IV doses of UC-MSCs in ELGA patients impacts (improves or worsens) all-cause in-hospital mortality, respiratory outcomes, complications of prematurity, neurodevelopment at 24 months corrected age, and long-term health outcomes.

### Study Population

Our study will target ELGA infants who remain ventilator-dependent with elevated oxygen requirements, aiming to identify those at greatest risk of developing BPD and most likely to benefit from early intervention.

#### a. Inclusion Criteria

An infant can participate once a parent or substitute decision maker has provided written informed consent. The infant must meet <u>all</u> the following inclusion criteria:

− Gestational age less than 28 weeks.
− Post-natal age between 4 and 14 days.
− On mechanical ventilation with FiO2 ≥ 0.3. An intubated patient with any of the following ventilation modes: conventional, high-frequency ventilation (oscillation or jet), and with the requirement of FiO2 ≥ 0.3 for at least 12 hours over the last 24 hours.

#### b. Exclusion Criteria

− Congenital anomalies, including genetic and chromosomal syndromes, major congenital anomalies, and inborn errors of metabolism.
− Shock, defined by hemodynamic instability with impaired end-organ perfusion (metabolic acidosis with increased lactate and/or decreased urine output) and requirements for fluid bolus, inotrope, or vasopressor medication.
− Severe sepsis, defined by hemodynamic instability (requiring at least one fluid bolus) and a positive blood or cerebrospinal fluid culture.
− Pneumothorax with chest tube in place.
− Severe pulmonary hemorrhage, defined as active pulmonary hemorrhage associated with at least one of the following: hemodynamic instability and/or need for blood product transfusion.
− Extubation planned within the next 24 hours.
− Not expected to survive (redirection of care or moribund).

### Enrollment process and randomization

The enrollment process involves identifying potential participants at NICUs and confirming their eligibility through health records and physician assessment. Substitute decision makers are approached for consent, with the aid of a dedicated animated informational video. Additional information for parents and families is accessible on the study webpage. Written consent is obtained using an ethics-approved form. The Ottawa Methods Centre at the Ottawa Hospital Research Institute will generate a computer-generated randomization list. Participants will be randomized in a 1:1 ratio using permuted random variable block sizes of 4 and 6 to avoid significant group imbalances. The Research Electronic Data Cap (REDCap) portal (35), based at the Women and Children’s Health Research Institute (WCHRI), will be used to confirm study eligibility and generate treatment allocation. We expect to have a 48–72-hour period between randomization and the delivery of the cell product for the first injection due to the cell manufacturing process and shipping. Therefore, there will be a re-evaluation of the exclusion criteria before administration and if there is a safety concern, administration will not go forward. The different steps of the enrollment process and randomization are presented in Supplementary Figure 1.

### Intervention Group

Participants allocated to the intervention group will receive a weekly (every 7 days ± 1 day) IV administration of 10×10^6 cells/kg of fresh human allogenic UC-MSC for 3 weeks.

Compared to our Phase 1 trial (Helping Underdeveloped Lungs with Cells Phase 1 Trial (HULC-1) (NCT04255147)), our Phase 2 trial will utilize a fresh cell product and multiple doses. Table 1 outlines the rationale behind the choices made for our intervention group, including cell tissue source (36–38), route of administration (23,29,34,39), multiple injections(19,34,40–42), and the use of a fresh cell product (43–47). Additional information on the cell manufacturing process and the cell product is provided as supplementary material. The cell product’s characteristics and manufacturing will be reported in accordance with clinical reporting guidelines for MSC trials (48).

**Table 1.** Rationale for the intervention group (UC-MSC)

| Component | Rationale | Supporting Evidence |
| --- | --- | --- |
| UC-MSc Source | UC-MSCs offer advantages over other MSC sources | • Readily available at birth from discarded tissue<br>• Higher initial cell yields (36)<br>• Superior anti-inflammatory, neuroprotective, and pro-angiogenic properties in vitro (36, 37)<br>• Greater effect on alveolarization and inflammation in BPD model compared to BM-MSCs (38) |
| IV Administration | Chosen over the intratracheal route for efficiency and safety | • IV MSCs retained in lungs via “first capillary filter” (39)<br>• Systematic review: IV route had greater effect on alveolarization (23)<br>• Good safety profile in adults and ELGA preterm infants (29, 34, HULC1 Unpublished, NCT04255147) |
| Multiple Doses | Multidose strategy aligns with BPD pathophysiology and enhances efficacy | • In chronic lung injury; repeated MSC doses may prolong anti-inflammatory and reparative effects<br>• Limited lung retention of UC-MSCs (19, 40)<br>• Superior effects on alveolarization and vascularization (41)<br>• Enhanced anti-inflammatory effects in preterm infants (34, 42)<br>• Improved outcomes in adults with ARDS (CIRCA-19 trial, NCT04400032)<br>• No dose-limiting toxicity in ELGA infants (34) |
| Fresh Cell Product | Fresh MSCs may be more effective than thawed cells | • Thawed MSCs show reduced immunosuppressive activity (43, 44)<br>• Improved clinical response with fresh MSCs (45)<br>• Immunosuppressive properties restored after 48h culture rescue (43)<br>• Darvadstrocel (FDA-approved MSC product) is a fresh preparation (46, 47) |

Numbers in brackets in the “supporting evidence” column refer to the references at the end of the manuscript.

The UC-MSC solution (5 milliliters) will be infused through a peripheral intravenous catheter using a syringe pump over 15 minutes, and an additional volume of normal saline will be infused over 15 minutes to flush the IV tubing dead space.

Even though clinical data in adult (29) and preterm (49–51) populations suggest a favorable safety profile for MSC, potential risks associated with IV administration of UC-MSC have been identified, and monitoring and risk-mitigation strategies have been defined and are presented in Table 2. In addition, adverse events will be managed in accordance with our study-specific infusion reaction protocol (supplementary material).

**Table 2.** Monitoring and risk-mitigation strategies to prevent potential risks related to UC-MSC IV administration.

| Potential risks related to IV UC-MSC * | Standard of care Monitoring in the NICU | Study specific risk mitigation strategies |
| --- | --- | --- |
| Infusional toxicity - fever | Infants admitted to the neonatal intensive care unit are continuously monitored with non-invasive devices for: heart rate, respiratory rate, blood oxygen saturation, temperature, blood pressure, and ventilator settings. | -Recording of vitals<br>-Refer to the study “Recommended Management for UC-MSC IV Infusion Reaction” document for guidance on actions to be taken (supplementary material) |
| Infusional toxicity - non-fever |  |  |
| Thrombo-embolic event |  | -Use of a filter on the IV line<br>-IV infusion over 15 minutes and saline flush post UC-MSC infusion over 15 minutes |
| Infection |  | NICU standard of care (monitoring of vitals) would enable early recognition and treatment for sepsis. |
| Coagulation |  | -Drug substance measured for tissue factor activity (ability to initiate thrombin formation). Only cells with low levels will be used.<br>-NICU standard of care monitoring for coagulopathy which could require bloodwork. |

\* Adapted from Thompson et al., E Clinical Med 2020 (29).
IV: intravenous, UC-MSC: umbilical cord-derived mesenchymal stromal cell, NICU: neonatal intensive care unit.

### Control Group

A placebo with IV infusion of the cell vehicle was initially considered. However, repeated peripheral IV line insertions solely for normal saline administration raised serious concerns regarding parental acceptance and potential impact on enrollment. We surveyed parents, who revealed that almost half were unlikely to participate in a study requiring peripheral IV insertions for normal saline infusion (unpublished data). Therefore, participants in the control group will receive a sham procedure weekly for 3 weeks to maintain double blinding without compromising enrollment. Patients randomized to the control group will not undergo IV catheter insertion; instead, we will simulate the procedure (IV catheter insertion and cell product infusion), and a bandage will be applied to the hypothetical insertion site. The use of sham procedure to maintain the blinding has been successfully applied in randomized controlled trials evaluating procedural intervention in preterm infants (52,53).

### Blinding

To maintain blinding of parents and healthcare providers to the study procedure (UC-MSC injection vs. sham), we will work with a blinded and an unblinded research team and implement the following protocol. The cell product or sham material will be delivered to the bedside in a sealed research box. The procedure will then be performed behind a privacy screen by the unblinded team (a nurse and/or physician). The individuals included in the blinded team will sign a non-disclosure agreement and will not participate in any other study activities, including data collection (except for collecting the vitals during and after the procedure) or adverse event reporting, neither in the clinical care of the participant. To assess the effectiveness of blinding, bedside staff will complete a questionnaire after each procedure, indicating their role (e.g., nurse, physician, respiratory therapist), their guess regarding the procedure performed (Intervention, Control, or Unknown), and their rationale.

### Outcomes

#### a. Primary outcome: Ventilation-Free Days Accounting for Mortality

The primary outcome of this study is the number of mechanical ventilation-free days (VFDs), measured at 120 days post-randomization and adjusted for mortality. VFDs will be calculated as follows:

**VFD = 0 days,** if the patient dies within 120 days or remains ventilated beyond that point.
**VFD = 120 minus days on mechanical ventilation,** if the patient is successfully weaned and survives within the 120-day window.

The primary outcome will be measured at 120 days post-randomization to allow the youngest potential participant to reach term-corrected age. In addition, according to Dassios et al., more than 99% of surviving patients born below 28 weeks should be off mechanical ventilation by postnatal day 100 (54). The 7-day free of mechanical ventilation difference was deemed by the study neonatologists as a clinically important difference needed to be observed to move to a Phase 3 trial. This outcome was selected by our research team based on its clinical relevance and ability to capture both duration of ventilation and survival as follows:

- **Clinical Relevance:** VFD is a primary outcome frequently used in adults and pediatric ARDS clinical trials (55). In preterm infants, duration of mechanical ventilation is associated with increased risk of BPD (56,57), intraventricular hemorrhage, and worse neurological development (58,59). Prolonged mechanical ventilation is also associated with a higher risk of death, requirement for home oxygen, and discharge on ventilation. Shortening mechanical ventilation is clinically and economically meaningful, as it reduces the level of care and potential painful procedures associated with it. Mechanical ventilation is also a key factor contributing to parental stress in the NICU (60–62).
- **Feasibility:** Ventilation-free days accounting for death is an appropriate composite outcome if the intervention is likely to reduce both ventilator duration and mortality (55). MSC’s pleiotropic effects support the hypothesis that UC-MSC administration in ELGA preterm infants might decrease the incidence of complications of prematurity, leading to a decrease in overall mortality.

#### b. Secondary outcomes

The secondary outcomes aim to assess the safety of IV administration of human allogeneic UC-MSC, the impact of these UC-MSC on the respiratory status, main complications of prematurity, and neurodevelopment at 24 months corrected age in our ELGA preterm infants study population. These secondary outcomes are fully represented in Table 3.

**Table 3.**
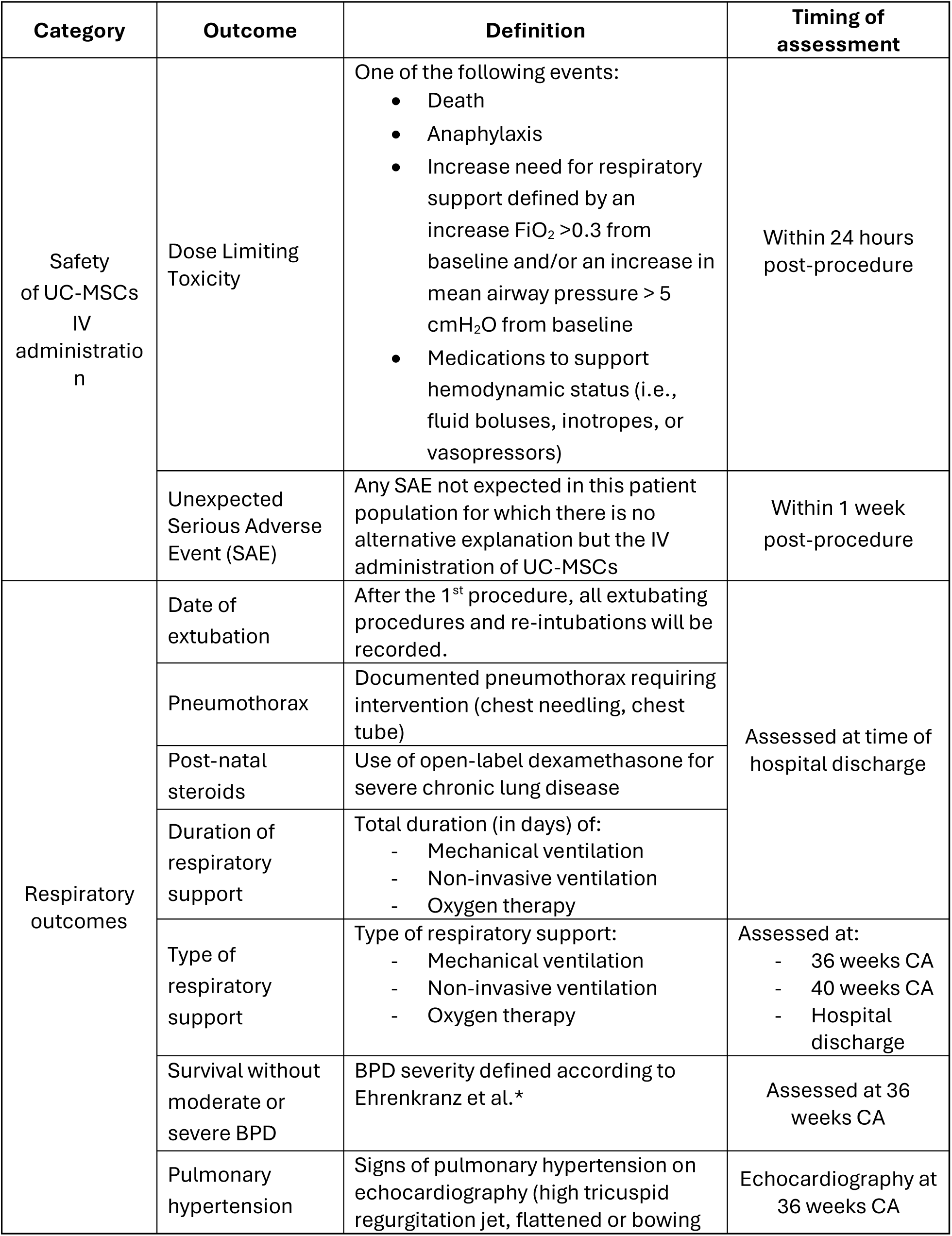
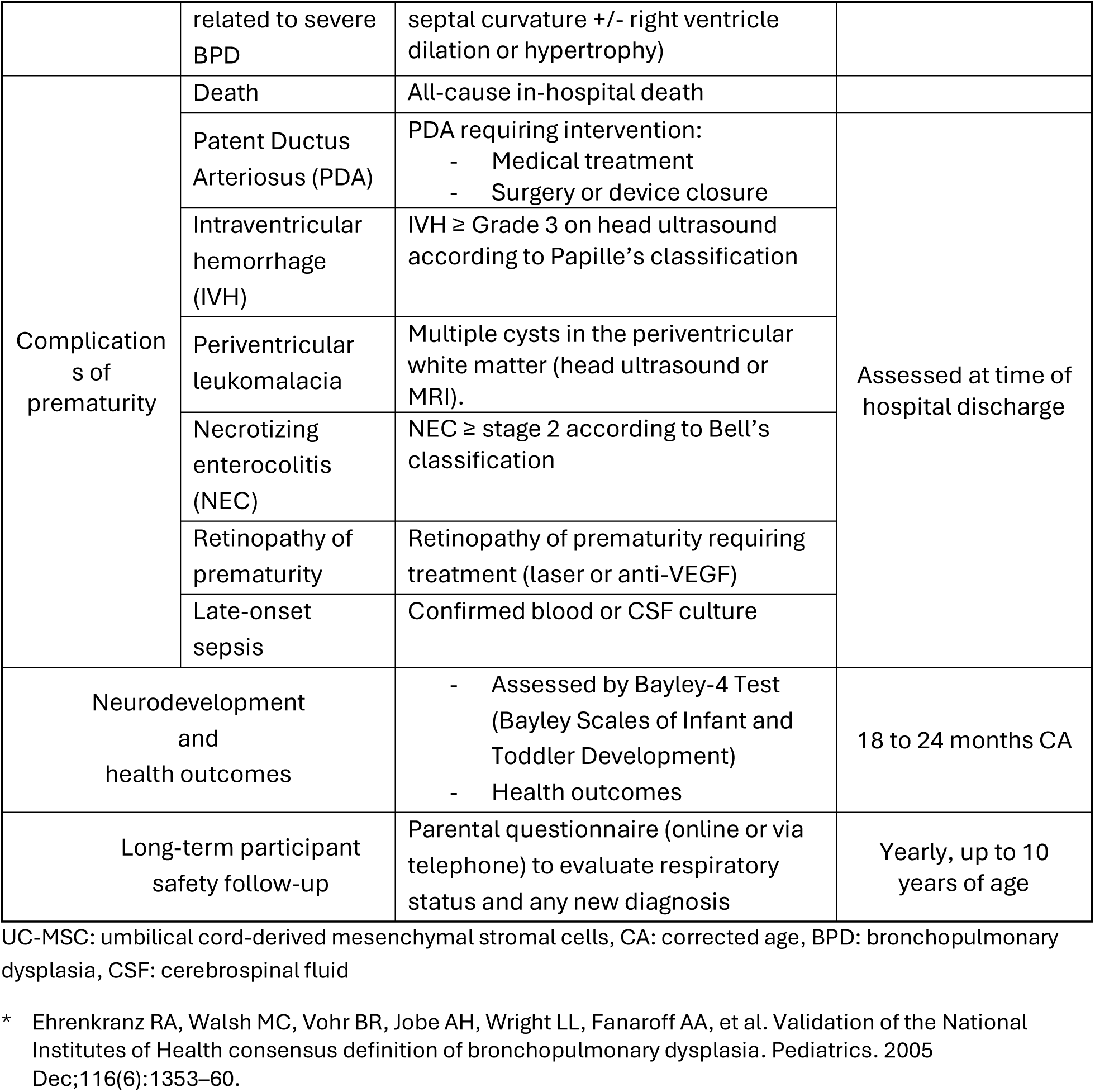
Study Secondary Outcomes.

### Sample Size Calculation and Data Analysis

A priori sample size calculation determined that 168 participants (84 in each study group) will be enrolled to achieve 80% power with an alpha of 0.05. This calculation accounts for a non-parametric distribution and aims to detect an increase of 7 days of mechanical ventilation-free days. The standard deviation of ventilator-free days for neonates <28 weeks is estimated to be 15, based on extrapolation from a large US study. The sample size has been inflated by 15% (Lehmann method) (63) to account for the non-parametric distribution. The primary outcome will be analyzed using the Mann-Whitney U Test. Median difference and 95% confidence intervals will be calculated. An adjusted analysis using quantile regression accounting for center, sex, gestational age, birth weight, and postnatal day of inclusion will be conducted. Secondary dichotomous efficacy outcomes will be analyzed with unadjusted Chi-Square tests and log-binomial logistic regression to produce unadjusted and adjusted relative risks and 95% confidence intervals. Continuous outcomes will be compared using appropriate parametric or nonparametric tests and generalized linear models.

### Study monitoring

For our trial, the Data and Safety Monitoring Committee (DSMC) will be composed of four members with expertise in cell therapies, in methodology and NICU clinical trials. The members of the DSMC are not affiliated with the Ottawa Hospital Research Institute and will declare any conflicts of interest if applicable. The DSMC members will act in an advisory capacity to the Qualified Investigator, Dr. Bernard Thébaud, to monitor patient safety. The DSMC will ensure that study participants will not be exposed to unnecessary or unreasonable risks and that the study will be conducted according to the highest scientific and ethical standards. The DSMC will determine if the study is safe to continue or if the study should be terminated.

The DSMC will review SAE as they are reported. They will meet to review data after each 10^th^ study participant (completion of study dosing/sham) procedure up to the 50^th^ study participant enrolled and subsequently after each 25^th^ study participant (completion of study dosing /sham) procedure until study completion.

### Ethics and dissemination Ethics

Ethics approval was obtained from the Ottawa Health Research Network Research Ethics Board (OHRN-REB #20250378-O1T) and Clinical Trials Ontario (CTO #5391).

### Registration and data availability

This clinical trial was registered on Clinicaltrials.gov on July 10, 2025 (NCT 07058025).

### Dissemination

This study adheres to the Ottawa Hospital Research Institute Publication Policy and is registered on ClinicalTrials.gov, with its protocol published. Results of this study will be shared on ClinicalTrials.Gov and presented at major national and international conferences and published in a scientific journal according to the reporting guidelines for clinical trials using MSC that were recently developed by our group (48). To ensure public understanding and engagement, patient partners will help create accessible materials for broader dissemination. Outreach efforts will include forums, student events, charities, medical associations, hospital foundations, and online webinars. Study participants will be able to access results via the study webpage (www.hulc-2.ca). Additionally, our lab’s website (http://xplorelab.ca) will provide regular updates and educational content to promote awareness of MSC biology and its potential to improve outcomes for preterm infants.

### Patient and public involvement

Two parent partners, as well as the Canadian Premature Babies Foundation (http://www.cpbf-fbpc.org/), are part of our research group to ensure that the preferences of future participants are considered. They were involved from inception in protocol development, reviewed all study documents for parents and families, including the consent form, provided feedback during the animated video and study website development, and will be further involved during each step of the study up to dissemination. They provided important insights on relevant variables and long-term outcomes for parents, the study design, and considering the acceptable ‘burden’ of additional study procedures. Their input and insights are crucial for developing more meaningful research for patients and increasing study acceptance among parents.

## Data Availability

This is the study protocol

## Authors’ contributions

LR: study design and protocol development, draft of the manuscript.

EF, CH, RG, LDS, VO, CCD: study design and protocol development. Revision of the manuscript.

DF, MR, MM, SH, SK, DC: Cell product documentation and revision of the manuscript. DF: study design, statistical analysis plan and revision of the manuscript.

BT: Funding, study design and protocol development, and revision of the manuscript.

---

**Supplementary Figure 1.**
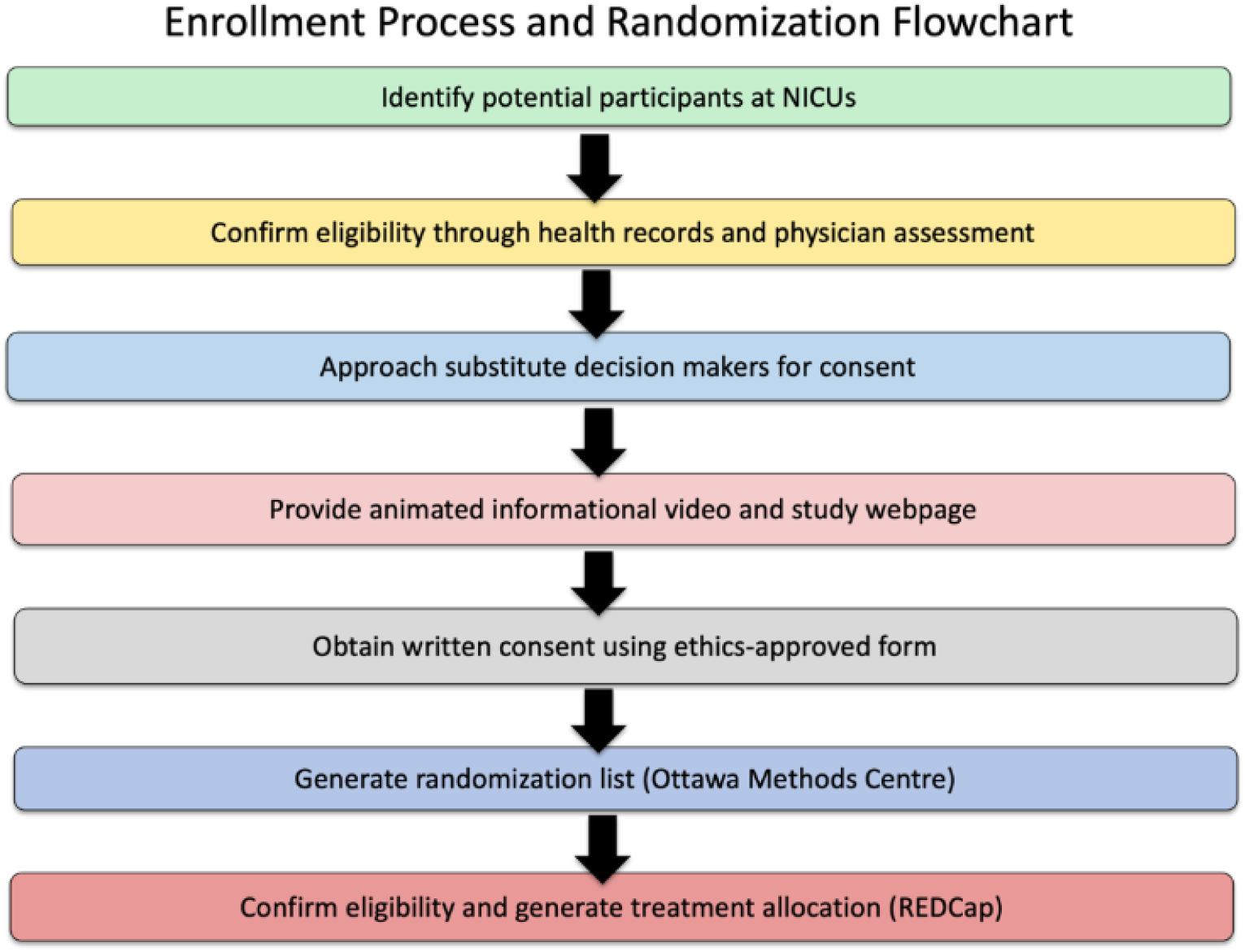
HULC-2 enrollment flowchart

## Supplementary material. HULC-2 Cell Manufacturing

### Donor Screening

Human umbilical cord tissue is obtained from healthy, term deliveries via C-section. In cases where the mothers fulfil pre-selection criteria, including uneventful pregnancy and absence of known chronic diseases, infections or genetic defects and who choose to donate their infant’s umbilical cord will undergo a rigorous screening protocol including a written sexual, social and medical history questionnaire, infectious disease testing and a physical examination on day of delivery. Infectious disease testing includes serological testing for HIV, hepatitis C virus (HCV), hepatitis B surface antigen, human T lymphotropic virus, cytomegalovirus (CMV) and syphilis, and nucleic acid testing for HIV, HCV and hepatitis B virus and is completed by the accredited/certified Institute of Medical Microbiology and Hygiene, Institute of Virology at the Technische Universität Dresden (TUD), Dresden, Germany.

### Tissue Collection

Umbilical cords are collected during scheduled C-sections performed at the Clinic for Obstetrics and Gynecology at the University Hospital Carl Gustav Carus Dresden, TUD, Dresden, Germany. Once the baby is safely delivered, the operating physician clamps and cuts the umbilical cord at a length of about 2 cm from the baby’s abdomen. After delivering the placenta with the remaining umbilical cord, the umbilical cord is milked carefully to remove cord blood under sterile conditions. After cutting the cord from the placenta, it is placed in a sterile, sealed, and clearly labelled transport container (400 mL re-sealable container, VWR International, USA) with 100 mL of Ringer’s lactate (Serumwerk Bernburg AG, Germany). The sealed transport container is picked up from the delivery room by the courier/employee of the GMP laboratory. The umbilical cord, together with the signed consent form and collection protocol, are transferred to the GMP laboratory for processing within 48 hours of delivery. A temperature monitoring device is put in place to ensure the package remains at 2 to 8°C during transit.

### Cell Isolation, expansion and cryopreservation

The cord tissue is transferred into the aseptic production area and disinfected with an iodine solution (Betaisodona; Mundipharma GmbH, Germany) followed by a washing procedure using Dulbecco’s Modified Eagle Medium (DMEM; Life Technologies Ltd, USA). Afterwards, the cord was mechanically and enzymatically dissociated using a mixture of collagenase (Nordmark GmbH, Germany), hyaluronidase (Riemser Pharma GmbH, Germany) and DNase (Hoffmann-LA Roche Ltd, Switzerland). Following further rinsing steps, the cell pellet was resuspended in complete culture medium (DMEM, 5 % Platelet lysate (PLBioScience GmbH, Germany) and 4mM Glutamax (L-alanyl-L-glutamine; Life Technologies Ltd, USA). Obtained cells were counted using the Nucleocounter NC-200 device (Chemometec Inc., USA) and seeded in T175 tissues culture flasks (Greiner Bio-One International GmbH, Germany) with complete culture medium and cultivated under hypoxic conditions (5% oxygen (O_2_), 5% carbon dioxide (CO_2_) at 37°C) for 5 to 7 days. After culture, umbilical cord tissue-derived mesenchymal stromal cells (UC-MSCs) were passaged using TrypLE (Life Technologies Ltd, USA) into HYPERFlasks (Corning Inc., USA) for further expansion. UC-MSCs are further cultured for an additional 5 to 7 days prior to harvesting and cryopreservation at 6 million cells/mL in freezing media (5% (w/v) Human Serum Albumin (Octalbin-5, Octapharma, Switzerland) and 10% (v/v) DMSO (WAK Chemie Medical GmbH, Germany). The cell suspension is aliquoted in 2mL per cryovial and formulated into a UC-MSC cell bank consisting of 200 – 300 vials.

### Product Release

UC-MSC cell bank is released for clinical use only after the following criteria are met:

- Cell viability is >80% as measured by Nucleocounter NC-200 at the time of cryopreservation
- UC-MSCs are positive (>95%) for CD73, CD90 and CD105 and negative (<2%) for CD14, CD34 and CD45. Cells are also negative (<10%) for HLA-DR
- Ability to form >10 colonies (CFU-F)
- Demonstrate enhanced Indoleamine 2,3-Dioxygenase (IDO) expression when challenged with interferon-gamma (ΔΔCt >10)
- UC-MSCs have endotoxin levels of <2 EU/mL
- UC-MSCs are free from microbial contamination after 14 days of culture in anaerobic and aerobic conditions
- Free from mycoplasma contamination with testing performed using the Sartorius ATMP Mycoplasma Detection Kit based on EP 2.6.7 and EP 2.6.27
- UC-MSCs have no major chromosomal abnormalities as per single nucleotide polymorphism-based genotyping performed at the accredited SYNLAB (Germany)
- Free from adventitious viral contaminants and negative for HBV, HCV and retroviruses with analysis performed at WuXi App tec (USA)

### Drug Product (DP) Preparation

After patient randomization to the cell treatment, cryovials from the qualified cell bank are thawed and diluted in complete media. The diluted UC-MSCs are seeded in T175 flasks (Corning, USA) and cultured for 48 to 120 hours in 5% O_2_, 5% CO_2_ at 37 °C. After 3 to 5 days in culture, cells are washed with PBS (Life Technologies, USA) and detached by treatment with protease (TrypLE, Life Technologies, USA) and subsequently diluted in equal volume of 5% HSA in 0.9% NaCl. The cell suspension is centrifuged, and the cell pellets are resuspended in 5% HSA-NaCl. UC-MSCs are filtered to form a single cell suspension and counted using Trypan Blue and a haemocytometer and the Drug Product is prepared at 3 million cells/mL.

To generate the patient specific dose of 10 million cells/kg pt weight, the DP is diluted by the addition of 5% HSA-NaCl. Five mL of the dose is drawn into a 10 mL sterile, single-use syringe (BD, USA) and capped with a sterile luer-lock cap. The sealed syringe is labelled with participant ID and expiry date/time then placed in a sterile sample bag, which is labelled as per Health Canada Division 5 Guidelines for labelling (C.05.011). This bag is placed in a second sterile bag and packaged into a validated and conditioned 2-8°C transport container with accountability log and temperature data logger. The transport container will be shipped to the site of cell administration by a courier service.

## HULC-2 Study Protocol Recommended Management for UC-MSC IV Infusion Reaction

This document was developed to provide guidance and can be adapted by the bedside team according to local policies and each clinical situation.

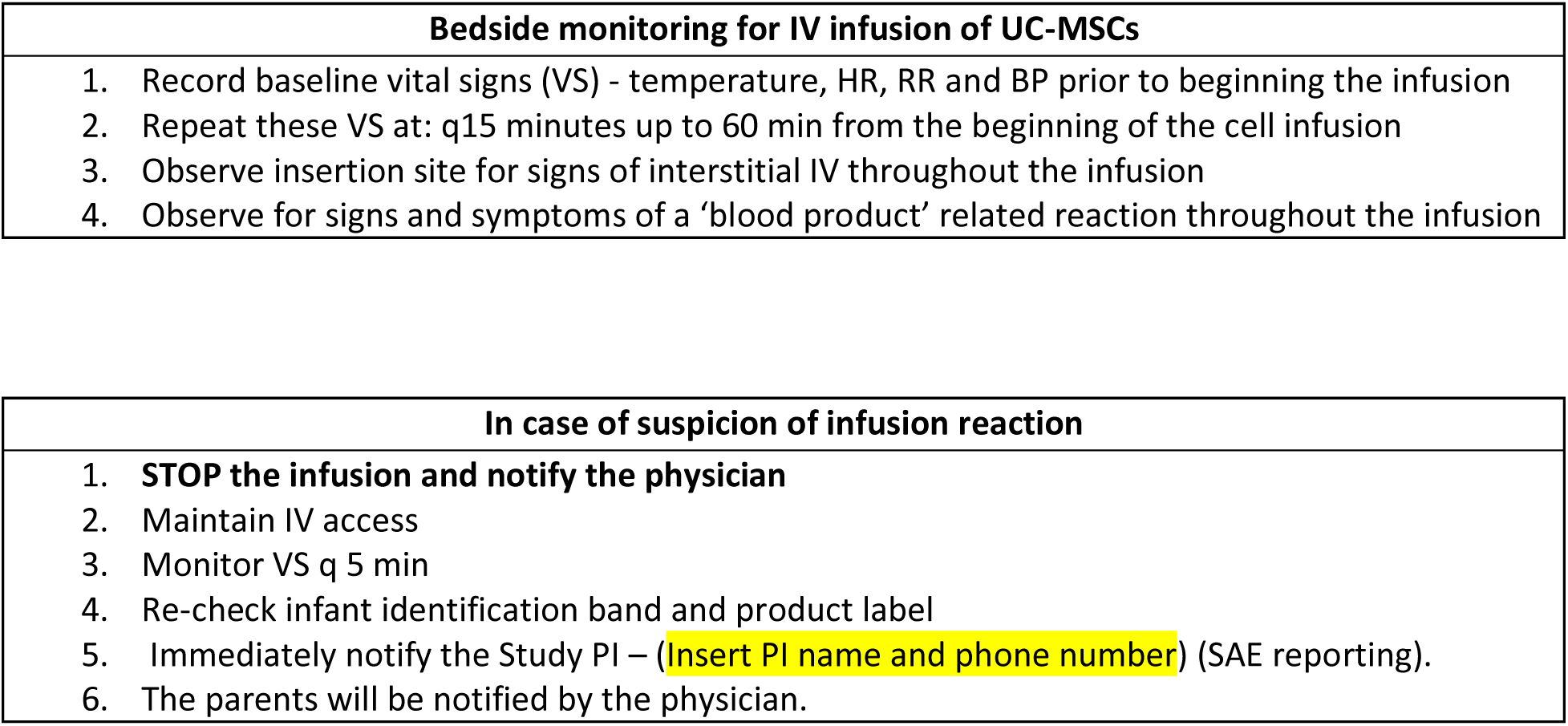

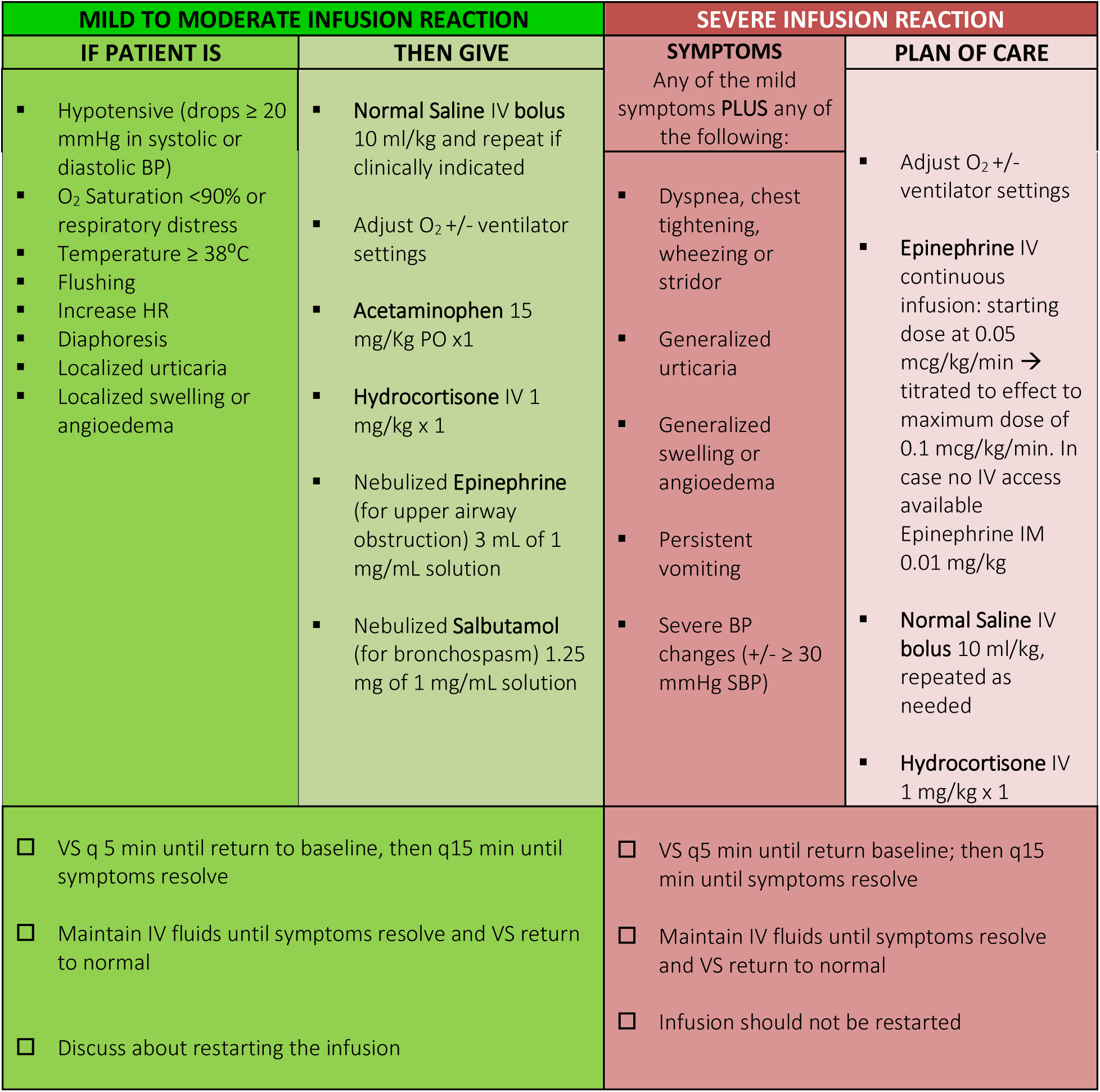

| MILD TO MODERATE INFUSION REACTION |  | SEVERE INFUSION REACTION |  |
| --- | --- | --- | --- |
| IF PATIENT IS | THEN GIVE | SYMPTOMS | PLAN OF CARE |
| ▪ Hypotensive (drops $\geq 20$ mmHg in systolic or diastolic BP)<br>▪ O<sub>2</sub> Saturation $<90\%$ or respiratory distress<br>▪ Temperature $\geq 38^{\circ}\text{C}$<br>▪ Flushing<br>▪ Increase HR<br>▪ Diaphoresis<br>▪ Localized urticaria<br>▪ Localized swelling or angioedema | ▪ **Normal Saline IV bolus** 10 ml/kg and repeat if clinically indicated<br>▪ Adjust O<sub>2</sub> +/- ventilator settings<br>▪ **Acetaminophen** 15 mg/Kg PO x1<br>▪ **Hydrocortisone IV** 1 mg/kg x 1<br>▪ Nebulized **Epinephrine** (for upper airway obstruction) 3 mL of 1 mg/mL solution<br>▪ Nebulized **Salbutamol** (for bronchospasm) 1.25 mg of 1 mg/mL solution | Any of the mild symptoms **PLUS** any of the following:<br>▪ Dyspnea, chest tightening, wheezing or stridor<br>▪ Generalized urticaria<br>▪ Generalized swelling or angioedema<br>▪ Persistent vomiting<br>▪ Severe BP changes (+/- $\geq 30$ mmHg SBP) | ▪ Adjust O<sub>2</sub> +/- ventilator settings<br>▪ **Epinephrine IV** continuous infusion: starting dose at 0.05 mcg/kg/min → titrated to effect to maximum dose of 0.1 mcg/kg/min. In case no IV access available Epinephrine IM 0.01 mg/kg<br>▪ **Normal Saline IV bolus** 10 ml/kg, repeated as needed<br>▪ **Hydrocortisone IV** 1 mg/kg x 1 |
| ☐ VS q 5 min until return to baseline, then q15 min until symptoms resolve<br>☐ Maintain IV fluids until symptoms resolve and VS return to normal<br>☐ Discuss about restarting the infusion |  | ☐ VS q5 min until return baseline; then q15 min until symptoms resolve<br>☐ Maintain IV fluids until symptoms resolve and VS return to normal<br>☐ Infusion should not be restarted |  |

**References**

- The Ottawa Hospital, Maternal, Newborn and gynecology service Standard Operating Procedure MNWH310 ‘Blood and blood products: Indication, administration and management’.

